# Association between Stage 1 Hypertension Defined by the 2017 ACC/AHA Hypertension Guideline and Cardiovascular Risk: A Large Cohort Study from the UK

**DOI:** 10.1101/2020.04.19.20071514

**Authors:** Fu-Rong Li, Xian-Bo Wu

## Abstract

**Importance:** The 2017 American College of Cardiology (ACC)/American Heart Association (AHA) hypertension (HTN) guideline lowered the threshold for HTN from 140/90 mmHg to 130/80 mmHg for systolic/diastolic blood pressure (SBP/DBP), resulting in a newly defined stage 1 HTN with an SBP/DBP reading of 130–139/80–89 mmHg. Few studies have assessed the impact of the redefined HTN on cardiovascular outcomes among the UK population.

**Objective:** To examine the effects of the revised ACC/AHA stage 1 HTN blood pressure parameters on the prevalence of HTN and related cardiovascular disease (CVD) risk in a large UK population.

**Design:** Adult men and women from a national cohort study in the UK.

**Setting:** The UK Biobank Study.

**Participants:** A total of 470,625 adults (mean age 56 years) with available data on blood pressure (BP) and without a history of CVDs at baseline.

**Main outcome measures:** Incident composite CVD outcome.

**Methods:** Prospective CVD events were analysed for survival in relation to BP measures using Cox proportional hazards regression models, adjusting for potential confounders. The associations are described by hazard ratios (HRs) and 95% confidence intervals (CIs).

**Results:** By adopting the 2017 ACC/AHA HTN guideline, an additional 24.7% of the participants were classified as having ACC/AHA stage 1 HTN, which resulted in a prevalence of HTN of 75.1% at baseline. During a mean follow-up period of 8.1 years, ACC/AHA stage 1 HTN (130–139/80–89 mmHg) was associated with a significantly increased risk of CVD (HR 1.20; 95% CI 1.10–1.30) compared to the risk associated with a normal BP (<120/80 mmHg). The excess risk of CVD associated with ACC/AHA stage 1 HTN was mainly driven by myocardial infarction (HR 1.19; 95% CI 1.05–1.36) and haemorrhagic stroke (HR 1.40; 95% CI 1.08–1.81), rather than ischaemic stroke (HR 1.02; 95% CI 0.87–1.19) and CVD death (HR 1.07; 95% CI 0.90–1.26).

**Conclusions:** The adoption of the 2017 ACC/AHA guideline would lead to a dramatic increase in the prevalence of HTN in the UK Biobank cohort study. Evidence from the present national cohort study may support lowering the threshold for HTN from 140/90 mmHg to 130/80 mmHg in the UK.

## Introduction

Hypertension (HTN) remains the most important risk factor for cardiovascular disease (CVD) and the leading cause of morbidity and mortality globally ^1, 2^ In 2017, the American College of Cardiology and the American Heart Association (ACC/AHA) released a guideline for the prevention, detection, evaluation, and management of high blood pressure (BP) in adults ^3^. A significant change in this guideline was a reduction of the threshold for HTN from 140/90 mmHg to 130/80 mmHg for systolic/diastolic BP (SBP/DBP). This change would include individuals who meet the definition based on the previous guideline in addition to a new group of adults labelled as having “stage 1 HTN” (130–139/80–89 mmHg).

To date, the lower targets in the new HTN guideline have received criticism for labelling people at low risk as having HTN, many of whom do not have any clear evidence of benefitting from additional treatment and monitoring ^4, 5^. In fact, as the benefits and harms of lowering BP could not be fully determined from current evidence, the existing BP targets have not been reduced in the new guideline in Europe and the UK—in the 2018 European Society of Hypertension (ESH)/European Society of Cardiology (ESC) Guideline, the definition of HTN remained unchanged, with HTN defined as office BP >140/90 mmHg ^6^. Similar to the European guideline, the National Institute of Clinical Excellence (NICE) in the UK has also chosen to maintain the previous definition of HTN^7^.

Indeed, although the impact of the 2017 ACC/AHA guideline on HTN management has been explored in different countries ^8-12^, it is unknown whether this recommendation can be applied to the UK population—an careful investigation of the health impacts of lowering the HTN criteria is crucial to facilitate evidence-based guidelines. Currently, however, there is little information regarding the clinical impacts of the revision of the ACC/AHA HTN guideline in the general UK population. More specifically, to what extent the ACC/AHA definition of stage 1 HTN affects the number of patients with HTN and the corresponding CVD risk in the UK population is a critical question to answer.

On this basis, we used data collected in the UK Biobank Study, a large national cohort of UK participants, to explore the effects of the 2017 ACC/AHA guideline for the management of HTN by estimating the prevalence of HTN and related CVD risks associated with ACC/AHA stage 1 HTN.

## Methods

### Study Population

The UK Biobank, a national long-term prospective cohort, comprises approximately 500,000 participants aged 40 to 69 years who were registered with the National Health Service (NHS) in the UK. Between 2006 and 2010, participants who agreed to take part in the UK Biobank visited 1 of 22 assessment centres across England, Wales, and Scotland for baseline assessments ^13, 14^ The information collected from the participants encompassed a broad range of characteristics, including sociodemographic data, physical measurements, lifestyle and clinical factors. In the present study, we excluded 1,243 individuals with missing values for SBP or DBP, 29,329 individuals with CVD events (angina, myocardial infarction [MI] and stroke) at baseline and 1,240 individuals who were lost to follow-up. The final sample for analysis included 470,625 participants.

The UK Biobank study was approved by the North West Multi-Centre Research Ethics Committee, and all participants provided written informed consent to participate in the UK Biobank cohort. Details of the UK Biobank are available elsewhere ^15^.

### Assessment of Blood Pressure

Two BP measurements were taken while the participant was seated after 5 minutes of rest using an appropriate cuff and an Omron HEM-7015IT digital BP monitor ^16^. Mean SBP and DBP values were calculated from 2 automated (*n*=439,829) or 2 manual (*n*=30,796) BP measurements.

### Assessment of the Outcomes

The primary outcomes of this study were composite CVD events: nonfatal MI, nonfatal ischaemic stroke (IS), nonfatal haemorrhagic stroke (HS), and CVD death. The secondary outcomes were the above individual outcomes. Information on CVD events and the timing of events was collected from cumulative medical records of hospital diagnoses and certified death records. At the time of analysis, hospital admission data were available through March 29, 2017, and mortality data were available through February 12, 2018, for England and Wales and August 12, 2016, for Scotland; these dates were used as the end of follow-up. Follow-up time was calculated from the date of baseline recruitment to the date of diagnosis of the event, death, or the end of follow-up, whichever came first.

The International Classification of Diseases, tenth revision (ICD-10) codes were used in death records, while ICD-10 and International Classification of Diseases, ninth revision (ICD-9) codes were used in medical records. MI was defined as ICD-9 codes 410-414 and ICD-10 codes I20-I25. IS was defined as ICD-9 codes 433–434 and ICD-10 code I63; and HS was defined as ICD-9 codes 430-432 and ICD-10 codes I60-I62 ^17^ CVD death was defined as ICD-10 codes I00-I99.

### Assessment of Covariates

Data on baseline characteristics were collected with a touch-screen questionnaire. We included the following covariates in our analysis: sociodemographic information (age, sex, ethnicity, and education level), area-based socioeconomic status (Townsend score); lifestyle (smoking, dietary habits and physical activity), kidney function (estimated glomerular filtration rate, eGFR), and self-reported medical conditions (diabetes and long-standing illness; drugs to treat high cholesterol and hypertension). A non-stretchable tape was used to measure height, and the Tanita BC 418 MA body analyser was used to measure weight ^18^ Body mass index (BMI) was calculated as weight in kilograms divided by height in metres squared (kg/m^2^). Dietary habits were assessed through a 24-hour dietary recall questionnaire. The ion selective electrode method (AU5400 Analyzer, Beckman Coulter) was used to measure sodium levels in stored urine samples ^19^ Serum cystatin C was measured by a latex-enhanced immunoturbidimetric method on a Siemens ADVIA 1800 instrument ^20^ Physical activity was assessed by the International Physical Activity Questionnaire short form. We used the summed metabolic equivalents of energy (METs) per week for all activity in the analysis. Based on previous studies of the UK Biobank ^17^, the healthy diet score was calculated using the following favourable diet factors: red meat intake no more than three times each week; vegetable intake at least four tablespoons each day; fruit intake at least three pieces each day; fish intake at least four times each week; cereal intake at least five bowls each week; and urinary sodium concentration less than 68.3 mmol/L. Each favourable diet factor was assigned a score of 1 point, and the total diet score ranged from 0 to 6. eGFR was calculated by CKD-EPI using serum cystatin C equations as previously reported ^21^. Long-standing illness was measured using the following question: “Do you have any long-standing illness, disability or infirmity?”.

### Statistical Analysis

Continuous variables are described as the means and standard deviations (SDs), and categorical variables are described by numbers and percentages. Participants were stratified into 5 BP categories: 1) normal (SBP <120 mmHg and DBP <80 mmHg), 2) normal high (SBP 120 to 129 mmHg and DBP <80 mmHg), 3) ACC/AHA stage 1 HTN (SBP 130 to 139 mmHg or DBP 80 to 89 mmHg), 4) NICE stage 1 HTN (SBP 140 to 159 mmHg or DBP 90 to 99 mmHg, the definition of stage 1 HTN according to the NICE guideline), and 5) moderate or severe HTN (SBP ≥160 mmHg or DBP ≥100 mmHg). Participants with normal BP were used as the reference. For supplementary analysis, we combined the normal BP group and normal high group, resulting in a new normal BP category (<130/80 mmHg); we also repeated the analysis after generating a separate group consisted of participants taking antihypertensive medications.

We applied multivariable Cox regression models to estimate the hazard ratios (HRs) and 95% confidence intervals (CIs) for the outcomes of interest, adjusting for potential confounders that may be associated with both BP and CVD. Based on tests using Schoenfeld’s residuals, there was no evidence of violation of the proportional hazards assumption. Responses of “not known” or “prefer not to answer” to the covariates were combined into an ‘unknown’ category. Missing values accounted for < 2.5% of all the covariates, except for physical activity (19.6%) and education level (17.9%). Participants with missing values for any of the adjustment variables were assigned to a separate “unknown” category for the respective variable. Two models were used. Model 1 was adjusted for age. Model 2 was adjusted for age, sex, ethnicity (white, mixed, Asian or Asian British, black or black British, Chinese or other ethnic group), BMI (in quintiles), education level (college or university degree, A/AS levels or equivalent, O levels/GCSEs or equivalent, CSEs or equivalent, NVQ or HND or HNC or equivalent, or other professional qualifications), Townsend scores (in quintiles), smoking (current/previous/never), alcohol consumption (never, special occasions only, 1-3 times per month, 1 or 2 times per week, 3 or 4 times per week, or daily or almost daily), healthy diet score, summed METs per week for all physical activity (in quintiles), cholesterol-lowering medication use, antihypertensive medication use, history of diabetes, long-standing illness, and eGFR (in quintiles). We also used Cox models with penalized splines to present the associations between CVD outcomes and BP on a continuous scale ^22^, with 3 degrees of freedom for all analyses. Assuming the causality of the association between BP and CVD, the population-attributable risk (PAR) was used to estimate the proportion of CVD that can be attributed to certain BP stratum, using the prevalence and adjusted HR.

For the subgroup analysis, we examined the associations between BP categories and CVD stratified by age (<65 years or ≥65 years), sex, eGFR (<60 ml min^-1^/1.73 m^2^ or ≥60 ml min^-1^/1.73 m^2^), history of diabetes, and 10-year CVD risk (<10% or ≥10%, calculated by the ACC/AHA Pooled Cohort Equations ^23^). Several sensitivity analyses were performed for the primary outcome by excluding participants 1) who had CVD composite events in the first two years and 2) who had missing variables.

Analyses were conducted using Stata version 14.0 (College Station, Texas) and R version 3.4.2 (R foundation for Statistical Computing). A *P*-value <0.05 was considered statistically significant.

## Results

### Study Population and Outcomes

The sample of 470,625 individuals comprised 208,231 men and 262,394 women. The age of these participants ranged from 37 to 73 years, with a median age of 56 years at baseline. Compared with those with normal BP, participants meeting the criteria for ACC/AHA stage 1 hypertension were older, more likely to be male, white, and obese; they also tended to have a higher level of physical activity, take antihypertensive medications and cholesterol-lowering medications, have diabetes, suffer from long-term illness, have a lower level of eGFR and a higher 10-year CVD risk. Other characteristics are shown in Table 1. During a median follow-up time of 8.1 years, a total of 13,157 composite CVD events occurred, including 7,344 MIs; 3,415 ISs; 1,118 HSs; and 2,971 CVD deaths.

**Table 1.** Characteristics of the study population according to different BP categories.

| Characteristics | Normal BP | Normal high | ACC/AHA stage 1 HTN | NICE stage 1 HTN | Moderate or severe HTN |
| --- | --- | --- | --- | --- | --- |
| No. of participants | 63,395 | 53,611 | 116,166 | 154,726 | 82,727 |
| SBP, mmHg | 111.5 (6.2) | 124.4 (2.9) | 131.3 (6.0) | 146.9 (7.0) | 169.7 (13.1) |
| DBP, mmHg | 69.4 (5.8) | 73.1 (4.7) | 80.5 (5.6) | 86.1 (7.5) | 94.2 (10.3) |
| Age, years | 52.1 (7.8) | 54.6 (8.1) | 55.0 (8.0) | 57.6 (7.7) | 59.7 (7.1) |
| Men | 15,687 (24.7) | 21,210 (40.0) | 51,613 (44.4) | 77,661 (50.2) | 42,060 (50.8) |
| White | 55,668 (87.8) | 48,242 (90.0) | 104,481 (89.9) | 141,308 (91.3) | 76,165 (92.1) |
| College or University degree | 25,683 (40.5) | 19,844 (37.0) | 39,637 (34.1) | 46,929 (30.3) | 22,456 (27.1) |
| Townsend score | -1.1 (3.2) | -1.3 (3.1) | -1.3 (3.1) | -1.4 (3.0) | -1.5 (3.0) |
| Body mass index, kg/m<sup>2</sup> | 25.0 (4.0) | 25.9 (4.1) | 27.3 (4.6) | 28.1 (4.8) | 28.5 (5.0) |
| Current smoker | 8,081 (12.8) | 6,031 (11.3) | 12,745 (11.0) | 14,684 (9.5) | 7,180 (8.7) |
| Alcohol use (once or twice a week) | 17,292 (27.3) | 14,321 (26.7) | 31,055 (26.7) | 39,285 (25.4) | 19,838 (24.0) |
| Healthy diet score | 3.0 (1.4) | 3.0 (1.4) | 2.9 (1.4) | 2.9 (1.4) | 2.9 (1.4) |
| Physical activity, METs/week | 2521.7 (2554.5) | 2679.4 (2680.1) | 2614.4 (2697.9) | 2695.0 (2756.2) | 2764.6 (2790.5) |
| Hypertensive medication use | 1,683 (2.7) | 2,304 (4.3) | 8,913 (7.7) | 20,222 (13.1) | 14,914 (18.0) |
| Cholesterol-lowering medication use | 3,848 (6.1) | 5,229 (9.8) | 13,872 (11.9) | 25,801 (16.7) | 14,959 (18.1) |
| Diabetes | 2,072 (3.3) | 2,587 (4.8) | 6,645 (5.7) | 11,419 (7.4) | 6,394 (7.7) |
| Long-term illness | 15,946 (25.2) | 14,438 (26.9) | 33,831 (29.1) | 48,120 (31.1) | 25,743 (31.1) |
| eGFR, ml min<sup>-1</sup>/1.73 m<sup>2</sup> | 94.2 (15.2) | 91.5 (15.6) | 89.9 (15.8) | 87.1 (15.7) | 84.8 (15.6) |
| 10-year CVD risk, % | 9.12 (21.8) | 10.4 (20.6) | 11.5 (21.0) | 14.3 (20.6) | 18.4 (20.8) |

BP, blood pressure; eGFR, estimated glomerular filtration rate.
Continuous variables were described as the means (stand deviation), and categorical variables were described as numbers (percentage).

### Prevalence of Hypertension

We examined the prevalence of BP measurements indicating HTN using two thresholds (≥130/80 mmHg and ≥140/90 mmHg) by the ACC/AHA and NICE guidelines. Based on the threshold of 140/90 mmHg, 50.5% of the participants had HTN. Using the threshold of 130/80 mmHg, an additional 24.7% of the sample was categorized as having ACC/AHA stage 1 HTN, resulting in a HTN prevalence rate of 75.1%. When stratified by age groups, applying the 130/80 mmHg threshold instead of the 140/90 mmHg threshold increased the prevalence of HTN from 28.9% to 58.4% (for ages <45), from 35.3% to 64.4% (for ages 45–49), from 43.7% to 71.3% (for ages 50–54), from 51.1% to 76.5% (for ages 55–59), from 59.6% to 81.7% (for ages 60–64), and from 68.2% to 86.7% (for ages >64) (Fig. 1A). SBP levels increased with age (Fig. 1B); however, DBP levels peaked among those between 55 and 59 years old (Fig. 1C).

**Figure 1.**
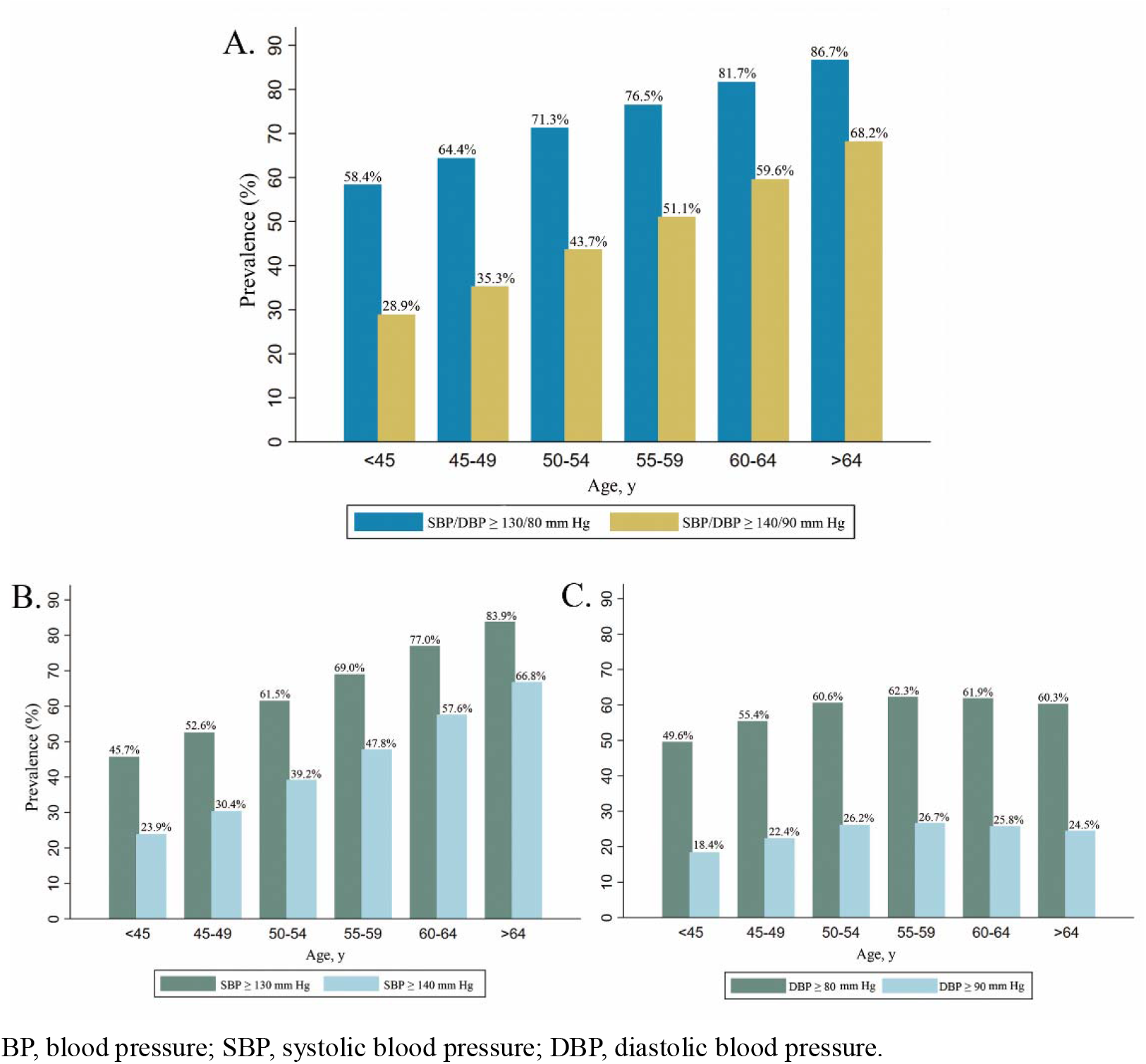
Relationship between age and BP measurements according to different thresholds.

### Associations between BP and CVD Outcomes

Compared with normal BP (<120/<80 mm Hg), ACC/AHA stage 1 HTN (130–139/80–89 mmHg) was significantly associated with an increased risk of CVD. Specifically, the fully adjusted HR associated with this BP category for the composite CVD outcome was 1.20 (95% CI 1.10–1.30); the risks of MI, IS, HS and CVD death were 1.28 (95% CI 1.14–1.43), 1.02 (95% CI 0.87–1.19), 1.40 (95% CI 1.08–1.81), and 1.07 (95% CI 0.91–1.26), respectively. For stage 1 HTN under the NICE guideline (140–159/90-99 mmHg), the corresponding risks were 1.44 (95% CI 1.34–1.56), 1.57 (95% CI 1.41–1.76), 1.28 (95% CI 1.11–1.48), 1.57 (95% CI 1.23–2.01) and 1.18 (95% CI 1.01–1.38), respectively (Fig. 2). Similar results were observed in the supplementary analysis, using a new normal BP group (<130/90 mmHg) as the reference, or generating a separate group of participants taking antihypertensive medications (Supplemental Fig. 1, Supplemental Fig. 2).

**Figure 2.**
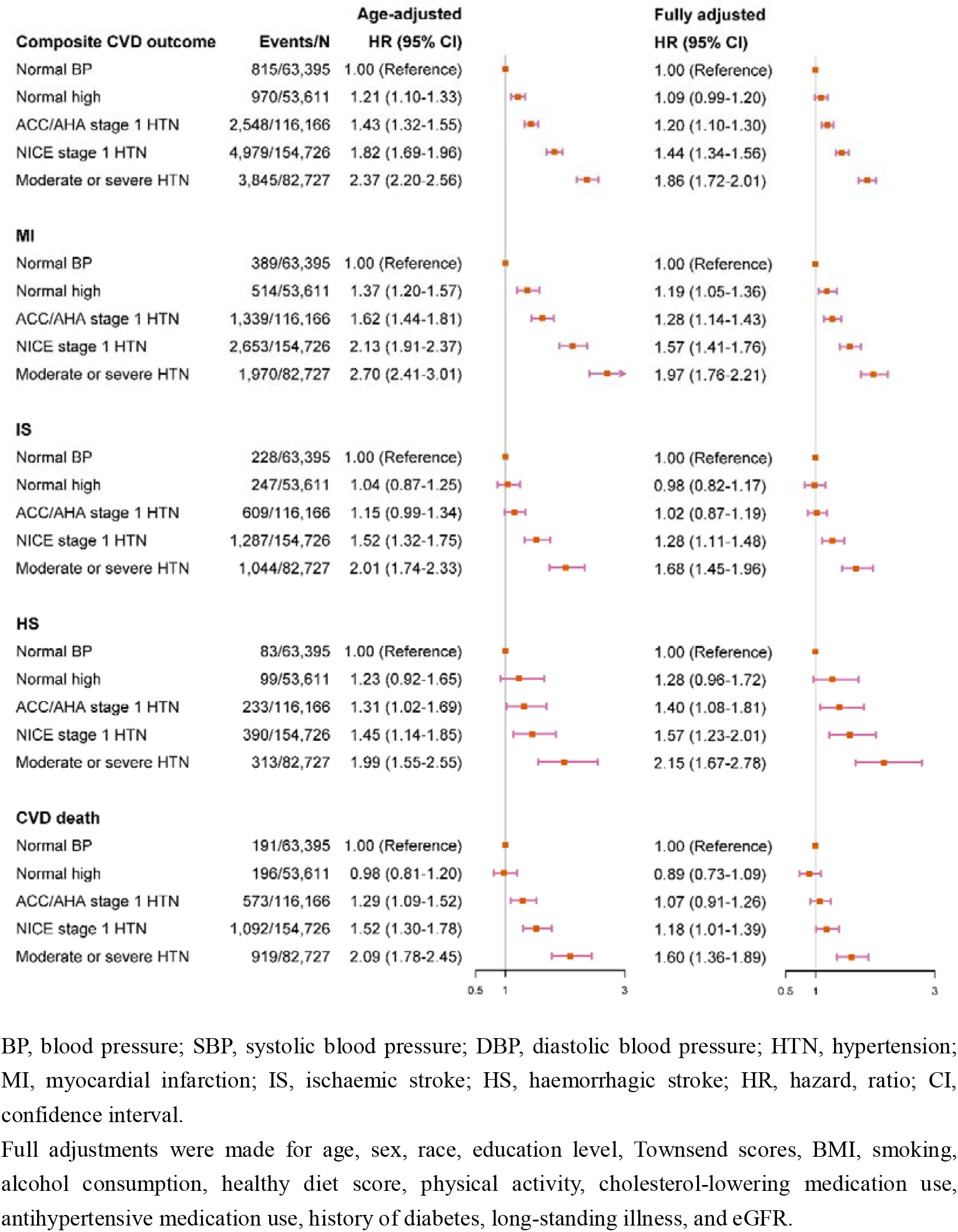
Hazard ratios and 95% CIs of CVD outcomes associated with different BP categories.

Compared with normal BP (<120/<80 mm Hg), the PAR for the CVD composite outcomes, MI, IS, HS and CVD death associated with ACC/AHA stage 1 HTN were 4.7%, 6.5%, 0.5%, 6.5% and 1.7%, respectively (Supplemental Fig. 3). A detailed study of the dose-response associations indicated that the links between BP measurements and the risk of CVD events increased largely monotonically across different CVD outcomes. However, J-shaped and U-shaped relationships were observed for the associations of both SBP and DBP with IS and CVD deaths (Supplemental Fig. 4 and Supplemental Fig. 5).

### Subgroup Analysis and Sensitivity Analysis

Subgroup analyses showed that the significant results between BP and CVD were mainly associated with younger adults aged <60 years and women (Table 2). When stratified by traditional risk factors, the significant results for ACC/AHA stage 1 HTN were seen only among those with eGFR ≥60 ml min^−1^/1.73 m^2^, without diabetes or with a 10-year CVD risk of <10% (Table 2). The primary results were similar after exclusion of participants who had CVD events in the first two years of follow-up or with missing variables (Supplemental Table 1).

**Table 2.** Hazard ratios and 95% CIs of CVD outcomes associated with different BP categories among various subgroups.

| Subgroups | Normal BP | Normal high | ACC/AHA stage 1 HTN | NICE stage 1 HTN | Moderate or severe HTN |
| --- | --- | --- | --- | --- | --- |
| Age |  |  |  |  |  |
| <60 years | 1.00 (Reference) | 1.10 (0.95-1.26) | 1.29 (1.15-1.44) | 1.61 (1.44-1.80) | 2.19 (1.95-2.47) |
| ≥60 years | 1.00 (Reference) | 1.03 (0.90-1.17) | 1.06 (0.95-1.19) | 1.24 (1.12-1.38) | 1.57 (1.41-1.75) |
| Sex |  |  |  |  |  |
| Men | 1.00 (Reference) | 0.89 (0.79-1.01) | 1.03 (0.92-1.14) | 1.23 (1.12-1.36) | 1.53 (1.38-1.70) |
| Women | 1.00 (Reference) | 1.34 (1.17-1.55) | 1.37 (1.21-1.55) | 1.64 (1.46-1.84) | 2.28 (2.01-2.58) |
| Diabetes |  |  |  |  |  |
| Yes | 1.00 (Reference) | 1.10 (0.84-1.44) | 1.10 (0.87-1.39) | 1.17 (0.93-1.46) | 1.42 (1.13-1.79) |
| No | 1.00 (Reference) | 1.07 (0.97-1.19) | 1.19 (1.09-1.30) | 1.47 (1.35-1.59) | 1.90 (1.75-2.07) |
| eGFR |  |  |  |  |  |
| <60 ml min<sup>-1</sup>/1.73 m<sup>2</sup> | 1.00 (Reference) | 0.76 (0.58-0.99) | 0.84 (0.66-1.05) | 0.89 (0.72-1.10) | 1.17 (0.95-1.44) |
| ≥60 ml min<sup>-1</sup>/1.73 m<sup>2</sup> | 1.00 (Reference) | 1.14 (1.03-1.26) | 1.26 (1.15-1.37) | 1.54 (1.42-1.67) | 1.98 (1.82-2.16) |
| 10-year CVD risk |  |  |  |  |  |
| <10% | 1.00 (Reference) | 1.09 (0.97-1.22) | 1.21 (1.09-1.33) | 1.45 (1.31-1.59) | 1.99 (1.78-2.22) |
| ≥10% | 1.00 (Reference) | 1.03 (0.88-1.20) | 1.08 (0.94-1.23) | 1.26 (1.11-1.44) | 1.57 (1.38-1.79) |

BP, blood pressure; HTN, hypertension; CVD, cardiovascular disease; MI, myocardial infarction; IS, ischaemic stroke; HS, haemorrhagic stroke; HR, hazard ratio; CI, confidence interval
Adjustments were made for age, sex, race, education level, Townsend scores, BMI, smoking, alcohol consumption, healthy diet score, physical activity, cholesterol-lowering medication use, antihypertensive medication use, history of diabetes, long-standing illness, and eGFR.

## Discussion

In the present national cohort study of the UK population, we found that the prevalence of HTN was 50.4% according to the NICE guideline. When adopting the 2017 ACC/AHA guideline, the prevalence of HTN was estimated to be 75.1%, with a 24.7% prevalence of newly defined HTN. During a median follow-up of 8.1 years, stage 1 HTN defined by the ACC/AHA guideline was associated with a significantly increased risk of CVD compared to the risk associated with normal BP and accounted for 4.7% of the CVD events among the UK population. The adverse effects associated with ACC/AHA stage 1 HTN were further supported by supplementary and sensitivity analyses.

Many studies of different countries and regions have shown that the 2017 ACC/AHA guideline resulted in a substantial impact on the prevalence of HTN ^8-12, 24^, although the proportion of HTN varies by cohort. The analysis of the NHANES showed that adopting the 2017 ACC/AHA HTN guideline in the US would categorize 63.0% of the population in the 45–75–year age group as having HTN, representing an increase in the prevalence of 26.8%. In China, the adoption of the new guideline would categorize 55.0% of the same age group as having HTN, an increase of 17.0% ^24^ In our study, we found that nearly three-quarters (75.1%) of the participants would be hypertensive based on the 2017 ACC/AHA HTN guideline, a higher prevalence relative to the data in the above nationwide survey of other populations. The variation in HTN prevalence may largely relate to differences in the sampling methods, methodology of assessing BP and population characteristics such as age, race/ethnicity, and cardio-metabolic status. A comprehensive exploration of the modifiable determinants of HTN in adults in the UK would be an important next step.

One of the main reasons for the NICE guidelines to maintain HTN definitions is that the results from randomized clinical trials (RCTs) do not firmly support additional benefits resulting from the new definitions. Guidelines such as the ACC/AHA guidelines have reduced their BP targets based on the results from RCTs including the US-based SPRINT and ACCORD trials ^25, 26^. However, both trials compared SBP treatment targets of 140 mmHg with more intensive targets of 120 mmHg. In addition, both trials included people who were at high risk for or already suffering from chronic conditions; therefore, the corresponding findings may not be simply generalized to the general UK population ^7^. Indeed, lowering the cardiovascular risk-based threshold will mean additional people requiring intervention, and treating the additional patients will require more public health resources. It has been estimated that the lower HTN criteria could result in as many as 450,000 more men and 270,000 more women in England being diagnosed with high BP and eligible for treatment ^27^ However, the public health impact of the HTN guidelines should mainly be interpreted in light of the expected beneficial effects on CVD outcomes ^28^, as it is well recognized that the early detection of HTN and subsequent intervention would slow BP progression, maintain vascular health, protect against organ damage, and ultimately lower CVD risk ^29^.

Of note is that the NICE guidelines do not recommend treating HTN for those with stage 1 HTN (140/90-159/99 mmHg) unless the individuals were thought to be of high risk (e.g., diabetes, eGFR<60 ml min^-1^/1.73 m^2^, or a 10-year CVD risk of ≥10%). In our data, however, high-risk populations such as those with diabetes or poorer kidney function were paradoxically not at risk for CVD at the BP strata of ACC/AHA stage 1 HTN or NICE stage 1 HTN. One of the explanations for these null associations might be related to bias from the higher comorbidity burden among the high-risk populations with low BP ^30, 31^; this was supported by the more linear relationship between BP and CVD after exclusion of those with high risk in our study (data not shown). Adequately designed trials from the UK are warranted to provide more robust evidence for the harms and benefits of lowering BP in high-risk individuals.

In our study, we found that the excess CVD risks associated with ACC/AHA stage 1 HTN were mainly driven by MI and HS, rather than IS and CVD death. Further, the U-shaped association of BP measures and CVD death in the present study may be of great importance for CVD prevention. On one hand, a markedly low BP is harmful and predictive of some adverse CVD outcomes ^31^; thus, caution must be taken when antihypertensive medication treatment is administered. On the other hand, rather than overemphasizing lowering BP, lifestyle changes should be the cornerstone of BP management, especially for individuals in the lower BP ranges. In fact, both the NICE and ACC/AHA guidelines advocate non-pharmacological therapy for patients in the early stage of HTN, unless they have a high-risk situation in which pharmacological treatment is additionally recommended ^6^.

We also observed that the newly defined HTN group was not associated with CVD risk in participants aged ≥60 years. This finding was consistent with previous meta-analyses of cohort studies showing that prehypertension was not linked to CVD risk among older adults ^32, 33^. Our study adds to the growing literature suggesting that the diagnosis of HTN and subsequent management of BP levels should be carried out with caution among older adults. The present study also found that the excessive CVD risk associated with ACC/AHA stage 1 HTN was observed only among women. Similarly, a study of 1.25 million UK patients also found that MI had a stronger association with SBP in women than in men ^34^. Overall, these findings may support tailoring recommendations for BP control for the primary prevention of CVD based on age and sex in the UK.

### Strengths and Limitations

The strengths of the present study include its very large sample size, the standardization of techniques to measure BP at baseline, and the detailed information on potential confounders. The linkage with death and hospital registries ensured complete information on the outcomes and minimized the number of participants who were lost to long-term follow-up.

Several limitations should be considered when interpreting the results of this study. First, BP was measured at a single visit, and the potential risk of bias due to “white-coat” effects should be taken into consideration, as a patient’s BP measured in a clinic or office may be higher than their ambulatory pressure. Therefore, the prevalence of HTN might be overestimated. Second, the study was not designed to collect a representative sample of the UK population, which limits the generalizability of the findings to the UK as a whole. However, as a nationwide cohort, with a risk factor profile comparable to those in other representative studies in the UK^35^, the results obtained from this study may reinforce the need for lowering the threshold for treating HTN in adults in the UK.

## Conclusions

The adoption of the 2017 ACC/AHA guideline would lead to a dramatic increase in the prevalence of HTN in the UK Biobank cohort study. Evidence from the present national cohort study may support lowering the threshold for HTN from 140/90 mmHg to 130/80 mmHg in the UK.

## Data Availability

The datasets and code used for the current study are available from the corresponding author on reasonable request. Example code used for the analysis is available on GitHub.

https://github.com/epimath/cm-dag

---

**Supplemental Table 1.** Sensitivity analysis for the associations between BP categories and CVD composite outcome.

|  | Hazard ratios (95% CIs) for SBP/DBP categories |  |  |  |  |
| --- | --- | --- | --- | --- | --- |
|  | Normal BP | Normal high | ACC/AHA stage 1 HTN | NICE stage 1 HTN | Moderate or severe HTN |
| Exclusion of those who had CVD within the first two years | 1.00 (Reference) | 1.14 (1.03-1.26) | 1.23 (1.12-1.34) | 1.49 (1.37-1.62) | 1.91 (1.76-2.09) |
| Exclusion of those with missing data | 1.00 (Reference) | 1.18 (1.04-1.34) | 1.29 (1.15-1.44) | 1.64 (1.47-1.82) | 2.15 (1.93-2.40) |

Adjustments were made for age, sex, race, education level, Townsend scores, BMI, smoking, alcohol consumption, healthy diet score, physical activity, cholesterol-lowering medication use, antihypertensive medication use, history of diabetes, long-standing illness, and eGFR.

**Supplemental Figure 1.**
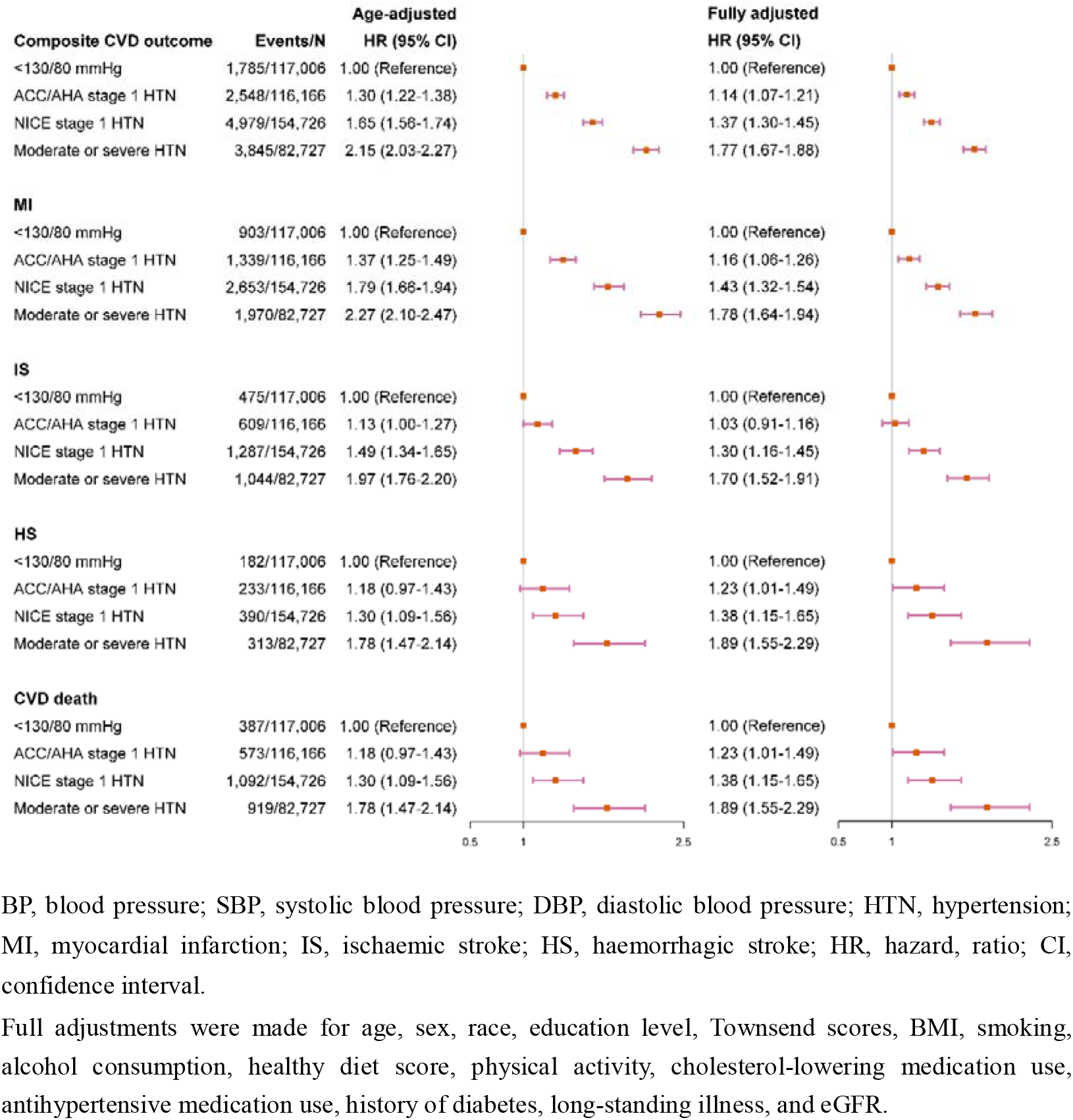
Hazard ratios and 95% CIs of CVD outcomes associated with BP categories, with BP of <130/80 mmHg as the reference.

**Supplemental Figure 2.**
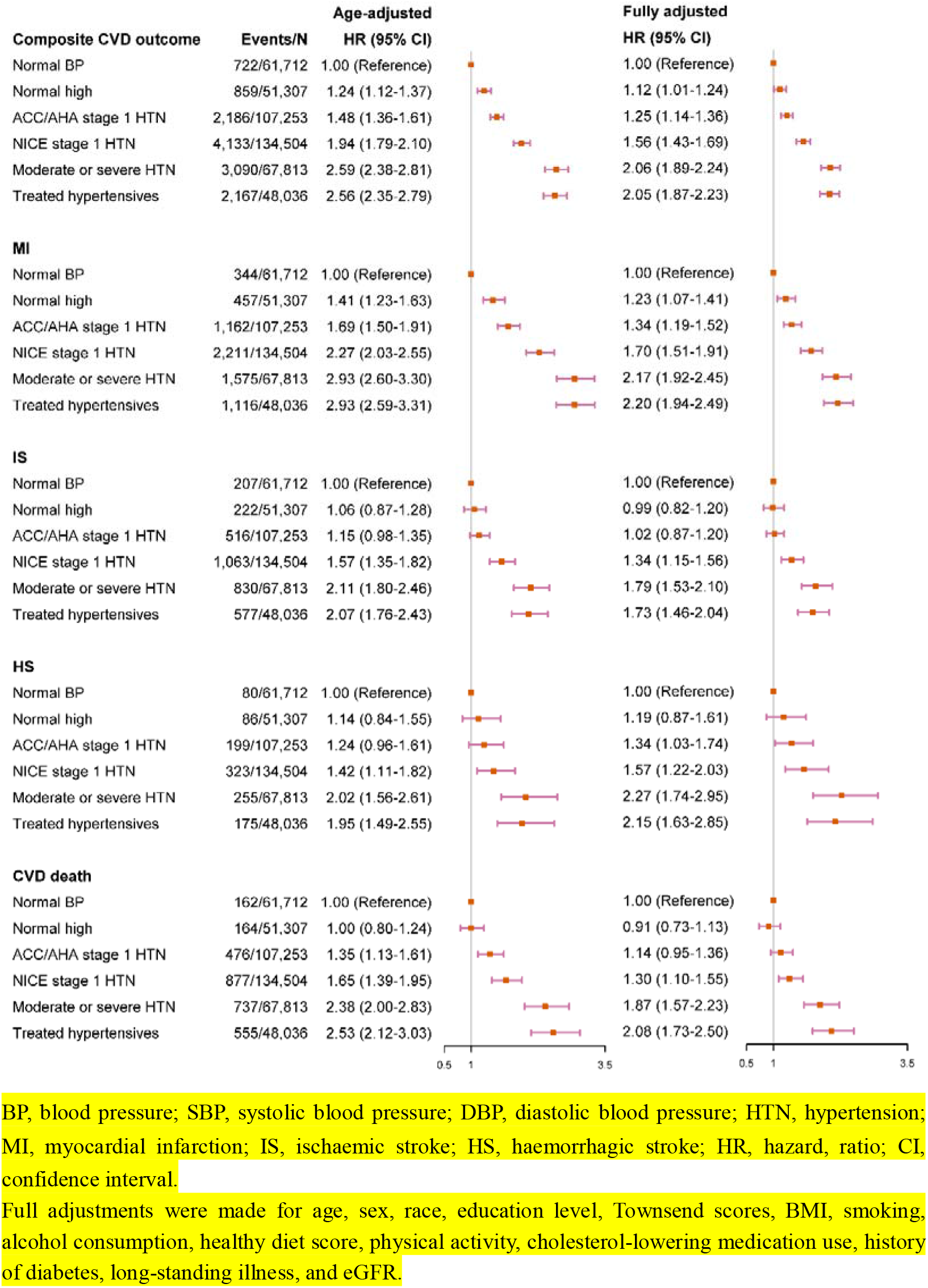
Hazard ratios and 95% CIs of CVD outcomes associated with newly defined BP categories, with those taking antihypertensive medications as a separate group.

**Supplemental Figure 3.**
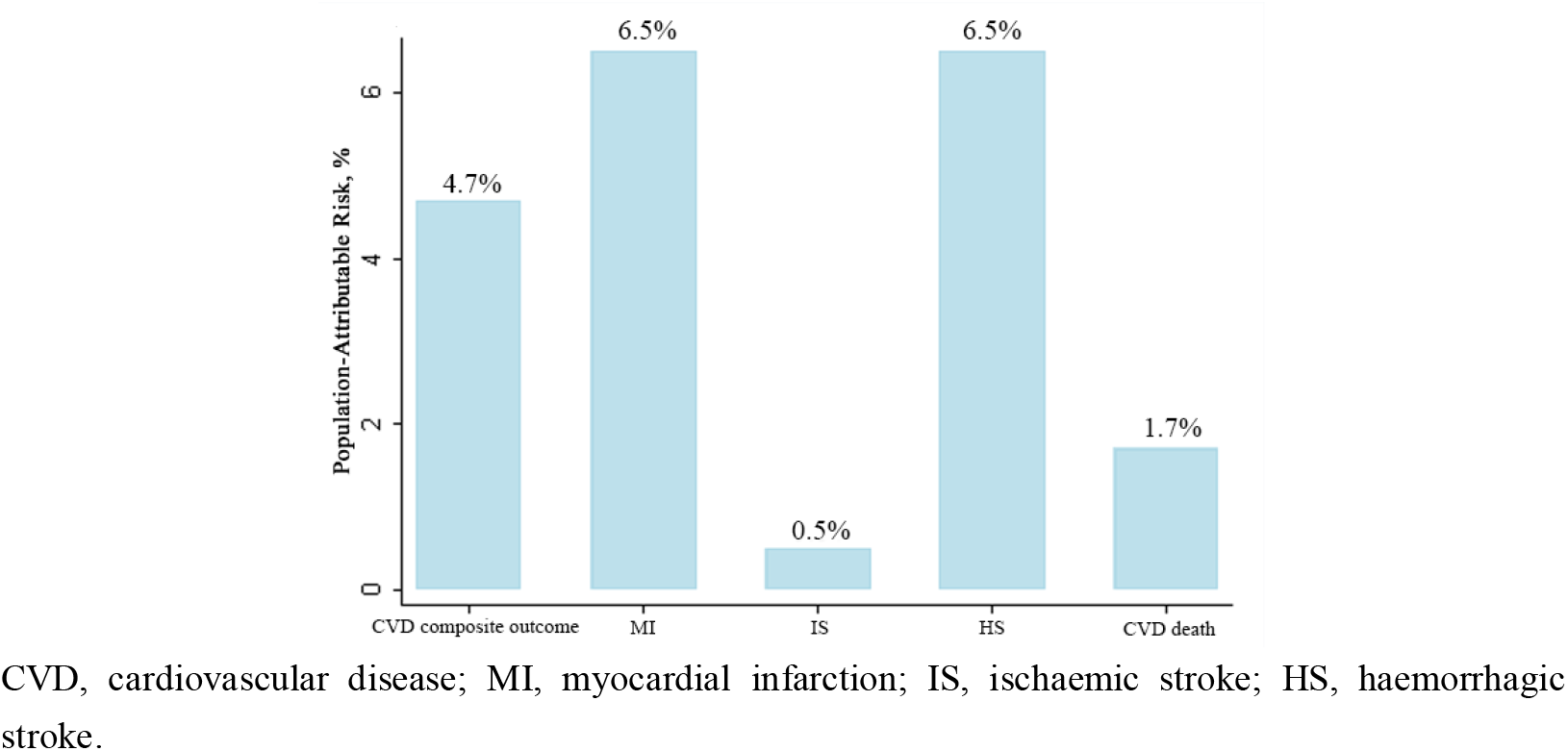
Population-attributable risk associated with ACC/AHA stage 1 hypertension for the whole population.

**Supplemental Figure 4.**
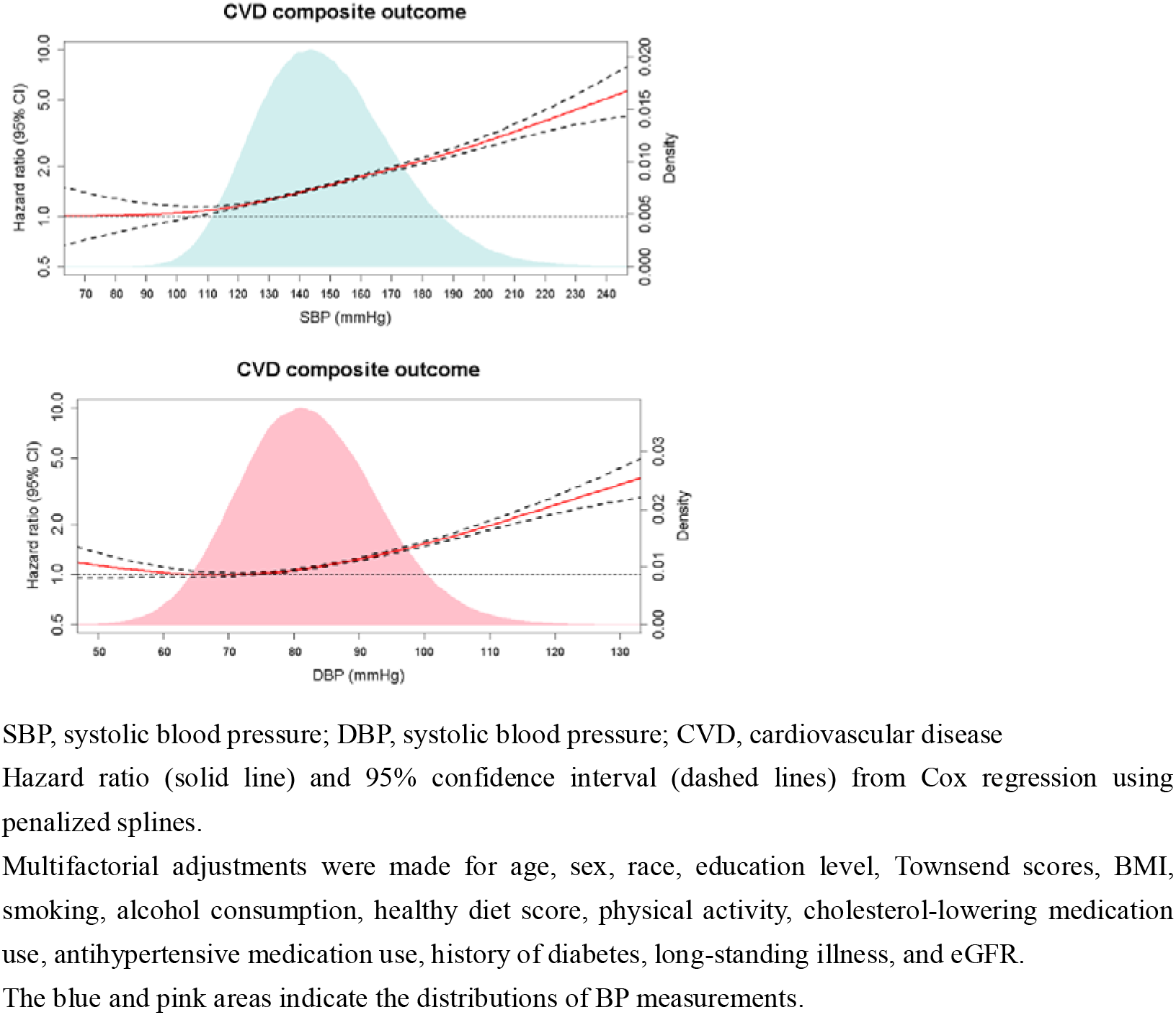
Blood pressure measurements on a continuous scale and risk of CVD composite outcome.

**Supplemental Figure 5.**
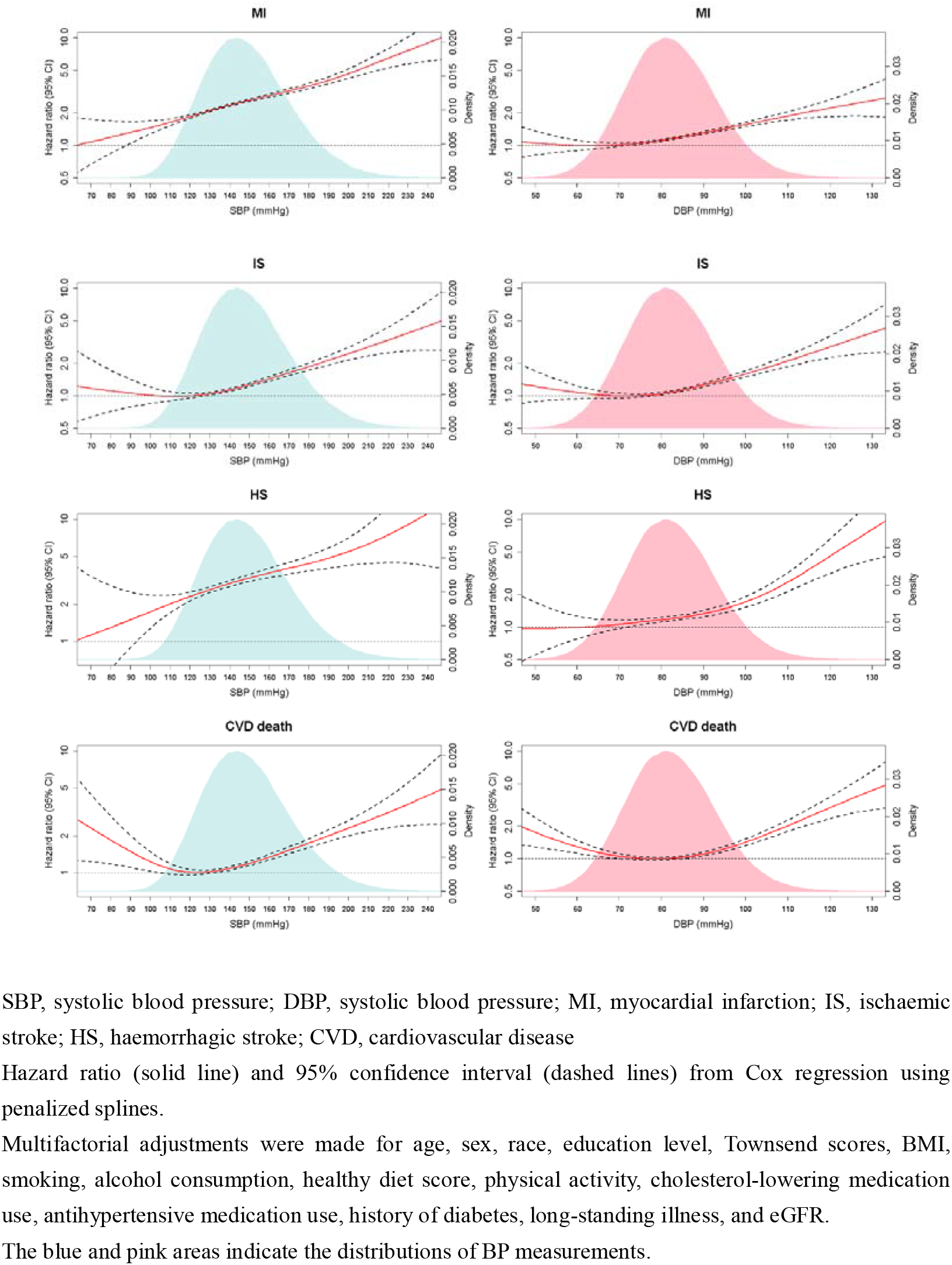
Blood pressure measurements on a continuous scale and risks of different CVD outcomes.

